# Genetic hearing impairment affects cochlear processing and, consequently, speech recognition in noise

**DOI:** 10.1101/2020.01.03.19015826

**Authors:** Cris Lanting, Ad Snik, Joop Leijendeckers, Arjan Bosman, Ronald Pennings

**Affiliations:** Department of Otorhinolaryngology, Radboud University Medical Centre, Donders Institute for Brain, Cognition and Behaviour, Nijmegen, The Netherlands

**Keywords:** Speech-in-noise, loudness growth, gap detection, frequency discrimination, otogenetics, hereditary hearing loss

## Abstract

The relation between speech recognition and hereditary hearing loss is not straightforward. Impaired cochlear processing of sound might be determined by underlying genetic defects. Data obtained in nine groups of patients with a specific type of genetic hearing loss were evaluated. For each group, the affected cochlear structure, or site-of-lesion, was determined based on previously published animal studies. Retrospectively obtained speech recognition scores in noise were related to several aspects of supra-threshold cochlear processing, as assessed by psychophysical measurements. The differences in speech perception in noise between these patient groups could be explained by these factors, and partially by the hypothesized affected structure of the cochlea, suggesting that speech recognition in noise was associated with genetics-related malfunctioning of the cochlea.

## Introduction

The relationship between speech recognition and the degree of sensorineural hearing loss is not straightforward. It was reported, for example, that patients with an average sensorineural hearing loss of 70 dB HL and adequate amplification obtained speech recognition scores that varied between 10 and 80% (Hoppe et al., 2014). Similar conclusions have been drawn for speech recognition in noise (Bosman and Smoorenburg, 1995; Vermeire et al., 2016). Ignoring a central neural processing deficit and other top-down influences such as cognitive factors as a cause (Humes et al., 2013; Kaandorp et al., 2016; Pronk et al., 2013; Stam et al., 2015), the poor relation between speech recognition and hearing impairment is supposedly related to variable degrees of deficient processing of speech by the impaired cochlea (Plomp, 1978; Plomp and Mimpen, 1979).

It was previously reported that patients with an autosomal dominant form of hereditary hearing loss (DFNA) type 2 and 9 (DFNA2 and DFNA9) with comparable high-frequency hearing impairment (measured in terms of the pure tone average across 1, 2 and 4 kHz, i.e. PTA_1,2,4 kHz_) had huge differences in speech recognition (Bom et al., 2001); DFNA9 patients with a PTA_1,2,4 kHz_ of 90 dB HL had an average phoneme score of 40% whereas this percentage for DFNA2 patients with the same degree of hearing loss was about 80%. Speech recognition-in-noise scores also seemed to be rather uniquely related to the underlying genetic type of hearing impairment (Leijendeckers et al., 2009). In line with these findings, we hypothesize that variation in speech recognition between patients is not primarily related to the degree of hearing impairment but more to the degree of impaired cochlear processing. The latter mainly depends on which part of the cochlea is affected; e.g. hair cells responsible for mechanotransduction (Gillespie and Müller, 2009; Peng et al., 2011), the stria vascularis and thus the endocochlaer potential (Lang et al., 2010), the tectorial membrane and the mechanical properties of the organ of Corti (Masaki et al., 2009). Over the last decade, we published the results on psychophysical and speech-in-noise tests obtained in nine different groups of hearing-impaired patients with a certain type of genetic hearing impairment (De Leenheer et al., 2004; Leijendeckers et al., 2009; Oonk et al., 2014, 2013; Plantinga et al., 2007; van Beelen et al., 2016, 2014, 2012; Weegerink et al., 2011) (see also Table 1). The present study uses the data from these publications to test our hypothesis. Furthermore, results from this study are important for the patient selection for, and evaluation of inner ear therapeutic studies and the further development of tests that characterise cochlear function and processing in more precise detail than current practice of pure tone audiometry and speech understanding.

**Table 1.** Included and excluded patients with genetic hearing impairment taken from previous studies. HDR stands for hypoparathyroidism, deafness, renal dysplasia syndrome; *: First author and year of publication.

| Type of genetic hearing loss | Gene | Reference * | n | Not in age range | Not in PTA range | Not in age AND PTA range | Final n |
| --- | --- | --- | --- | --- | --- | --- | --- |
| DFNA8/12 | *TECTA* | Plantinga, 2007 | 5 |  | 2 |  | 3 |
| DFNA10 | *EYA4* | Van Beelen, 2016 | 5 | 1 | 1 |  | 3 |
| DFNA13 | *COL11A2* | De Leenheer, 2004 | 14 | 2 | 3 | 1 | 8 |
| DFNA22 | *MYO6* | Oonk, 2013 | 3 |  |  |  | 3 |
| DFNB18B & DFNB84B | *OTOG* & *OTOGL* | Oonk, 2014 | 6 | 4 |  |  | 2 |
| Usher syndrome type 2a | *USH2A* | Leijendeckers, 2009 | 11 |  |  |  | 11 |
| Muckle Wells syndrome | *NLPR3* | Weegerink, 2011 | 5 |  | 2 | 2 | 1 |
| Non-ocular Stickler syndrome | *COL11A2* | Van Beelen, 2012 | 9 |  | 4 |  | 5 |
| HDR syndrome | *GATA3* | Van Beelen, 2014 | 3 |  |  |  | 3 |

### Patients and methods

In 2004, De Leenheer et al. introduced a test battery consisting of psychophysical (loudness perception, temporal and spectral processing) and speech recognition in noise tests to assess cochlear processing (De Leenheer et al., 2004). Over time, these tests have been used on nine different groups of patients with a specific type of genetic hearing loss (Table 1). Additionally, in some of these studies results have also been collected for normal hearing controls. The results from these previous studies were used in the present study to test the hypothesis that variation in speech recognition in noise between patients is related to the degree of impaired cochlear processing that in turn is likely caused by the underlying genetic disorder.

The test battery consisted of four different tests, performed in a standardised way in all patient groups. *Loudness growth* was measured with 0.5 kHz and 2 kHz tones. The best-fit curve through the loudness growth data was calculated and its slope was the primary outcome measure (Slope of the Loudness Growth, SLG). It was decided to present the slope relative to (divided by) the slope of normal hearing subjects. This means that a relative slope of 1 reflects a loudness growth similar to that of normal hearing subjects. In case of loudness recruitment, the relative slope is larger than 1 (Dillon, 2012). *Gap detection (GDT)* or the shortest perceived period of silence between two noise bursts was measured using band-filtered white noise with center frequencies of 0.5 kHz and 2 kHz. To obtain a relative measure, the smallest detectable gap is presented relative to (divided by) the norm value. A value of 1 means that the smallest gap detected is not different from the norm. *Difference limen for frequency (DLF)* was measured at 0.5 kHz and 2 kHz with frequency modulated tones. The lowest modulation frequency that was detected by the patient was taken as the DLF and it is presented relative to (divided by) the norm value. The critical signal-to-noise ratio (*S/N*) was measured by using the *speech recognition in noise test*, also refered to as the Plomp-test (Plomp, 1978; Plomp and Mimpen, 1979). These data are also presented relative to controls, i.e. the reported critical *S/N* values are compared to norm values (*S/N* patient – *S/N* controls) and this difference is called the Cochlear Distortion Factor (CDF) or D-factor (Plomp, 1978). If CDF is > 0, the patient has more difficulty to understand speech in noise than normal hearing individuals.

To investigate the present research hypothesis, we decided to homogenise the nine groups of patients and to only include data of patients aged between 18 and 70 years. These inclusion criteria were previously introduced and relate to presbyacusis as a factor that might interfere with hereditary hearing impairment (De Leenheer et al., 2004) and can thus be considered as an upper age limit, and to problems of compliance or reproducibility with the task observed in children and adolescents while performing some of the more complex subtests (Oonk et al., 2014), putting a lower limit on age. Furthermore, the degree of hearing loss was homogenised; the individual pure tone average (average hearing loss at 0.5, 1, 2 and 4 kHz, or PTA) had to be moderate to severe, between 30 dB HL and 75 dB HL. Table 1 presents numbers of selected and non-selected patients per study and Table 2 presents the mean age and mean PTA with their ranges. Figure 1 presents the mean audiogram per patient group; most audiograms are relatively flat or mildly sloping.

**Table 2.** Characteristics of the patient groups. HDR stands for hypoparathyroidism, deafness, renal dysplasia syndrome

| Type of genetic hearing loss | Age, in yrs.<br>(range) | PTA (0.5-4.0<br>kHz) in dB<br>HL (range) | Audibility-corrected<br>Cochlear distortion<br>factor (CDF) in dB<br>(range) |
| --- | --- | --- | --- |
| DFNA8/12 | 37 (27-45) | 40 (33-53) | 1.5 (0.9-2.4) |
| DFNA10 | 52 (31-65) | 62 (60-65) | 4.1 (4.6-6.0) |
| DFNA13 | 44 (34-63) | 39 (33-48) | 1.5 (0.6-2.6) |
| DFNA22 | 60 (53-66) | 41 (36-46) | 3.0 (2.5-4.1) |
| DFNB18B | 19 (18-20) | 43 | 3.6 (3.1-4.1) |
| Usher syndrome type 2a | 40 (28-59) | 52 (41-69) | 5.6 (3.8-9.2) |
| Muckle Wells syndrome | 21 | 60 | 6.6 |
| Non-ocular Stickler<br>syndrome | 58 (44-68) | 52 (46-58) | 4.1 (2.9-5.7) |
| HDR syndrome | 38 (25-56) | 54 (51-57) | 3.6 (2.0-4.8) |

**Figure 1.**
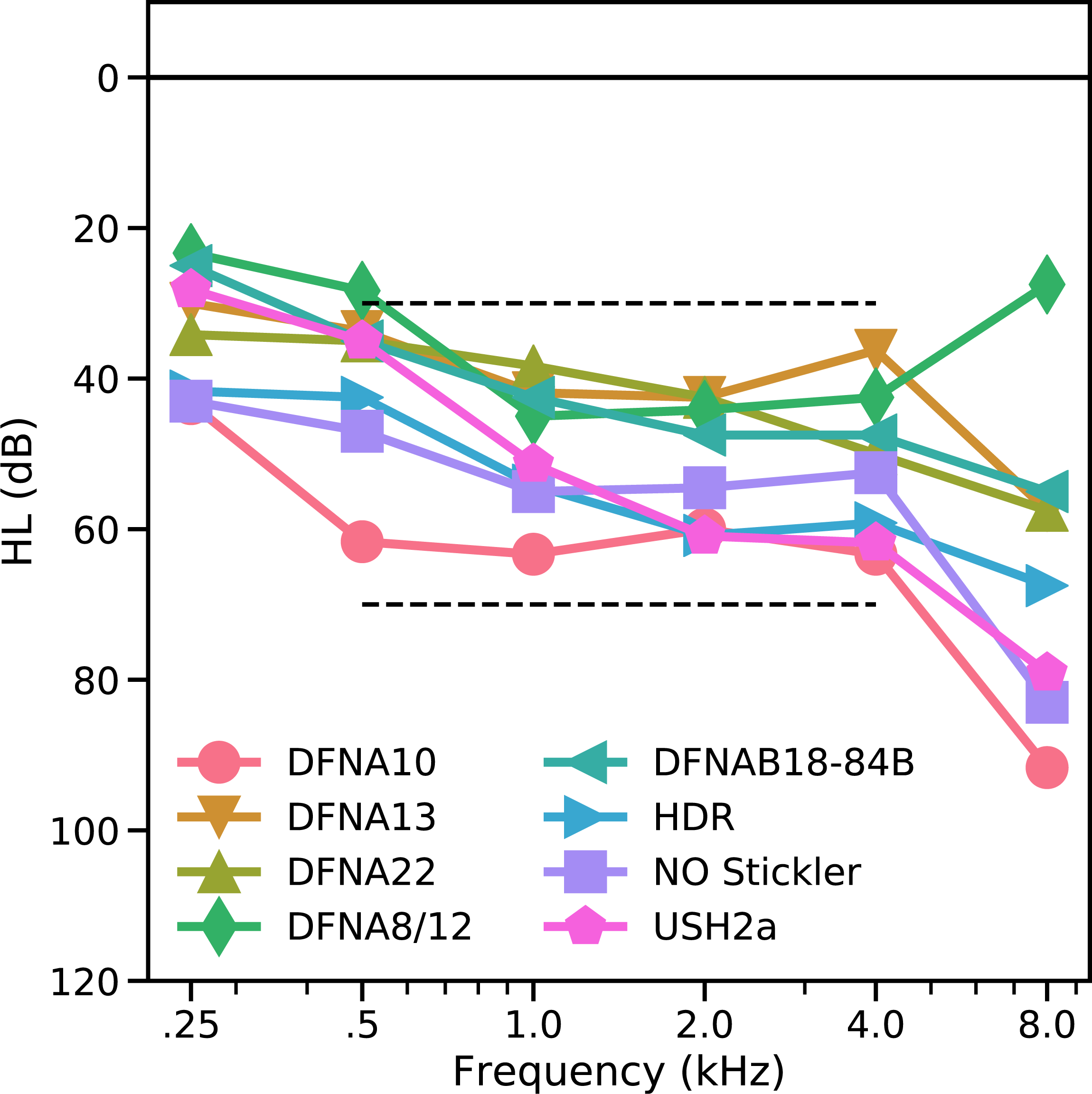
Mean audiogram of the patient groups. The circles and downward-pointing arrows refer respectively to the DFNA10 and DFNA13 patients; the upward-pointing arrow and the diamond symbols refer respectively to the DFNA22 and DFNA8/12 patients; the left- and right pointing arrows refer respectively to the DFNAB18-84B and HDR patients, and the squares and the hexagon symbols refer respectively to the NO Stickler and the USH2a patients.

For the Usher syndrome type 2a (USH2a) group, the DFNA10 group, the Non-Ocular (NO) Stickler syndrome group, and the HDR (Hypoparathyroidism, Deafness, Renal dysplasia syndrome) group, and in some individuals of other groups, a predominantly high-frequency hearing loss is seen (Fig 1). This might have consequences for the audibility of speech in the speech-in-noise test (Humes and Dubno, 2010). The *S/N* values are therefore corrected for inaudibility of speech in the higher frequencies, using the simplified method previously described (Killion and Christensen, 1998) and reviewed (Amlani et al., 2002). This correction is applied on individual data. For USH2a, the CDF of 8 of the 11 patients is corrected; the mean correction factor is 0.9 dB (range: 0.2-1.8 dB). The audibility-corrected CDF values are used for further analyses. For the HDR patients, corrections are found to be low (between 0 and 0.3 dB) and these were subsequently neglected. With the present inclusion criteria, the Muckle-Wells patient group comprised only one patient and was therefore excluded. The DLF test was not carried out in the group of DFNA10 patients because of logistic problems.

In retrospect, the speech-in-noise data are related to the loudness growth data, gap detection threshold and difference limen for frequency, using repeated regression analyses. In addition, for comparison with the literature, the speech-in-noise data are also related to the generic variables age and hearing loss. As the outcomes in speech-in-noise test depends primarily on processing of high frequency information (Bosman and Smoorenburg, 1995), the psychophysical data obtained at 2 kHz are used (i.e. the highest of the two frequencies tested). Two separate analyses have been performed: the first analysis related the speech-in-noise data to individual patients whereas the second analysis used the mean data per patient group.

## Results

Table 2, column 4 presents the mean CDF as calculated from the *S/N* outcomes of the patient groups together with its range, which is 2 dB or less in most patient groups, suggesting good reproducibility (Plomp and Mimpen, 1979). Audibility corrections of the CDF have been performed on individual patients. Figure 2 and Table 2 show that the CDF varies between patient groups; in DFNA8/12 and DFNA13 (Fig 1, first two box-plots and table 2, rows 1 and 3), its value is relatively close to 0 (the norm), which is indicative for (sub)normal speech recognition in noise. Poorest results are seen for the DFNA10 and USH2a patient groups (Fig 1, 7^th^ and 8^th^ boxplots and table 2, rows 2 and 6), even after audibility corrections.

**Figure 2.**
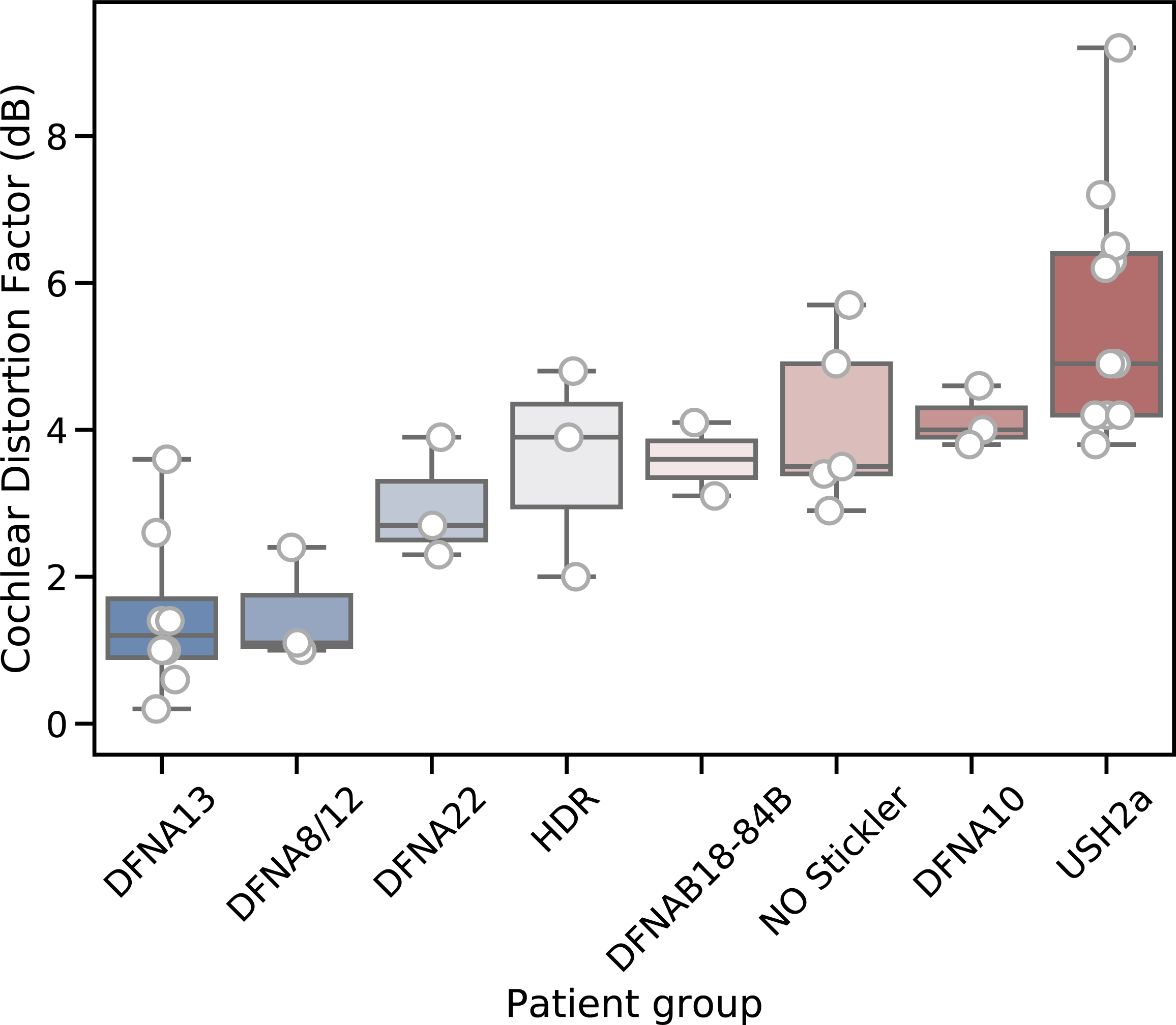
Boxplots of the eight patients groups, rank-ordered by the mean CDF for each group. The groups vary with respect to their mean CDF, but also with respect to the distribution within each group. The highest mean CDF (i.e. poorest speech understanding in noise) and the broadest distribution of individual patients’ CDF-values can be observed in the USH2a group.

Following the literature (Van Esch and Dreschler, 2015; Vermeire et al., 2016) the relation between speech in noise scores and the generic variables hearing loss (PTA) and age was also studied. Simple linear regression (ordinary least squares) of the CDF with variables PTA and age showed that the CDF was significantly related to the PTA (F (2,35) = 14.8, *p* < 0.001) with an r^2^ of 0.3. This relation shows that the CDF increased with 1.1 dB per 10 dB increase in hearing threshold (PTA). Interestingly, the intercept of the regression line does not cross the origin but instead shows an intercept of −2.0 dB (figure 3). By definition, the CDF is 0 for normal hearing subjects, thus seriously questioning the significance of the relation of CDF with PTA.

**Figure 3.**
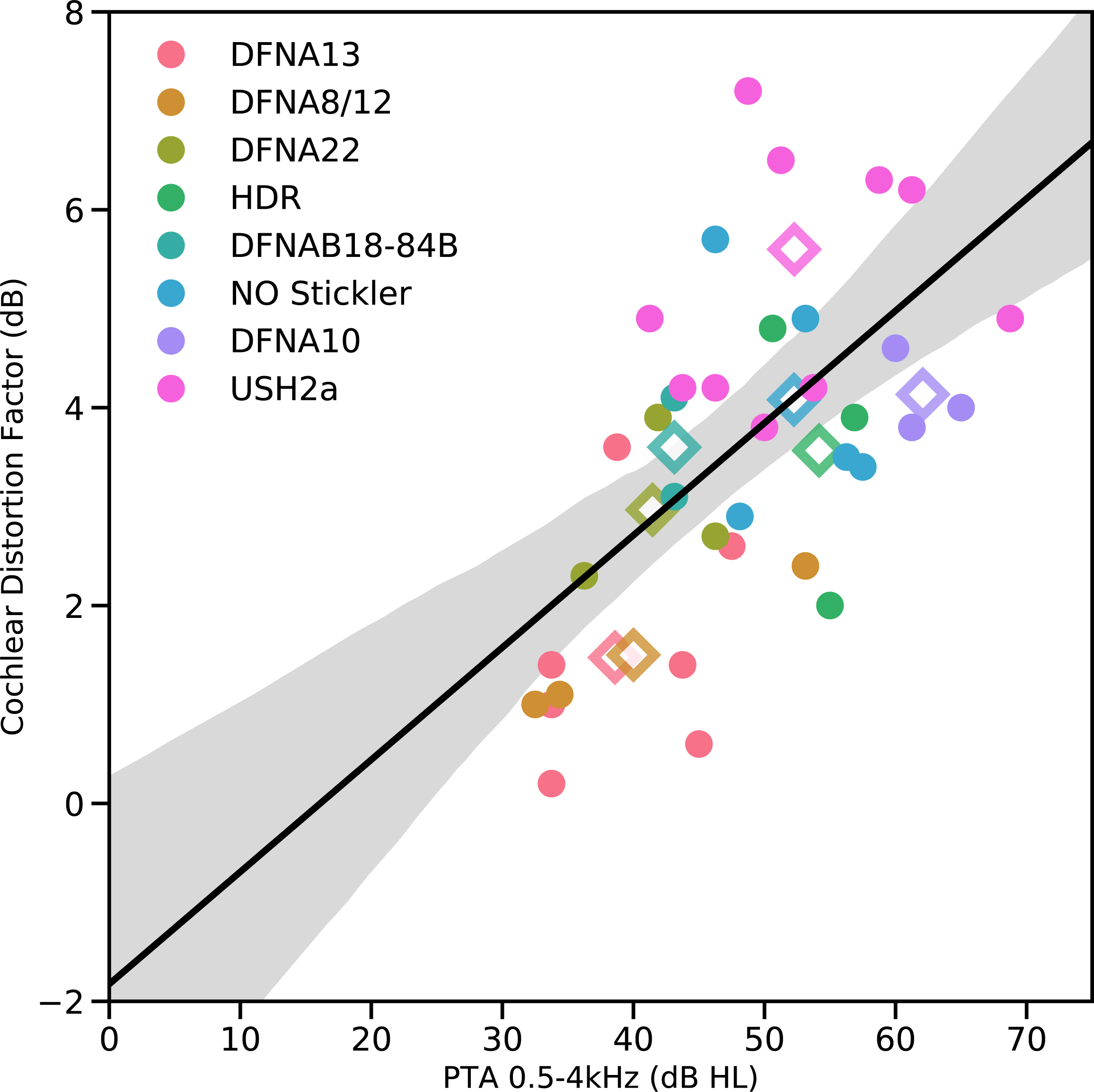
Cochlear distortion factor (CDF) as a function of the hearing loss (PTA) of the eight patient groups (individual data shown as circles where each patient group has a different color. The average for each patient group across PTA and CDF is indicated by a diamond-shaped symbol). The linear regression line presents the calculated best-fit curve and the 95% confidence interval of the fit.

Yet, despite this apparent relation, it can be observed that the CDF also varies with respect to the grouping variable *‘patient group’*. For some groups, such as the Usher2A patients, most patients (i.e. 7 out of the 10 patients that have a complete data-set) have a CDF that falls above the regression line and its 95%-confidence interval. In another group, such as the DFNA13 group, the majority (7 out of 8 patients) have a CDF that falls below the regression line. This indicates that although CDF and PTA seem related, the regression model with only PTA and age fails to capture most of the variance. Indeed, by adding the variable ‘patient group’ as a categorical variable, the model captures more explained variance from r^2^ = 0.30 (variables PTA and age) to r^2^ = 0.74 (variables ‘patient group’, PTA and age; model fit: F (12,22) = 5.1, *p* < 0.001). The variables PTA and age are not significant predictors of CDF anymore in this adjusted model. Adding the three psychophysical variables i) difference limen for frequency (*DLF*), ii) the relative slope of the loudness growth curve (*SLG*) and iii) the relative gap detection threshold (*GDT*) to the linear regression model does not increase the explained variance. Indeed, when looking at the individual factors, only the categorical variable for the patient group is significant F (7,22) = 4.77, *p* = 0.002). At the level of the individual patient, the psychophysical variables cannot predict the CDF.

Since the variation of CDF within the patient groups is substantial, linear regression analysis was also performed on the mean CDF data for each group and the three psychophysical variables. Linear regression analysis using the relative slope of the loudness growth curve (SLG) and the relative gap detection threshold (GDT) as variables raised the explained variance (r^2^) from 0.63 (only SLG) to 0.88 (group average model 1: both GDT and SLG; F(2,7) = 18.6, *p* = 0.004)). Adding the difference limen for frequency (DLF) instead of gap detection also improved the r^2^ from 0.87 to 0.98 (group average model 2: DLF and SLG; F(2,7) = 95, *p* < 0.001). These r^2^ values are obviously very high. The final equations for the two models are:

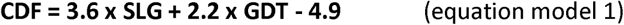

and

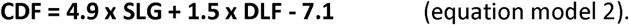

According to these two models, a normal hearing subject (SLG =1, GDT = 1, DLF=1) has a CDF factor of 0.9 dB and −0.7 dB, respectively for model 1 and model 2, thus close to the expected CDF of 0 (no cochlear distortion). At the level of the group the CDF could thus be predicted by the combination of the loudness growth (SLG) and either the relative gap detection threshold (GDT) or the relative difference limen for frequency (DLF). The fits are both significant, show a high degree of explained variance and show that the two models also hold for normal-hearing participants.

## Discussion

Using previously published data on different groups of patients with genetic hearing impairment, we have shown that cochlear distortions vary considerably between patient groups with different types of genetic hearing impairment and that the distortions neither relate to the severity of hearing impairment nor age. This variation in cochlear distortion between the various patient groups is suggestive of dysfunction at specific cochlear subsites (e.g. hair cells, tectorial membrane), or site-of-lesions, where some forms are more detrimental to understanding speech in noise than others. This in turn may provide an explanation for the often-reported poor relation between hearing loss (PTA) and speech recognition.

To further elaborate on this, we considered different types of cochlear hearing loss as proposed by Schuknecht and Gacek (Schuknecht and Gacek, 1993). Based on microscopic temporal bone studies of deceased patients with well-documented audiometric history, Schuknecht and Gacek distinguished four predominant types of cochlear hearing loss hypothesized to underly presbyacusis: 1. the sensory type (hair cell loss), 2. the strial type (atrophy of the stria vascularis), 3. the neural type (disproportional loss of auditory nerve cells) and 4. the cochlear conductive type (no significant loss of any tissue while the audiogram is mildly sloping). They also mention a mixed phenotype (i.e. a combination of the four types mentioned) as well as a remaining indeterminate type covering up to 25% of all presbyacusis cases. With growing knowledge in the field of genetic hearing loss and molecular biology of the inner ear, this classification system has been debated over the years (Lee, 2013; Ohlemiller, 2004). New insights based on fractional survival of hair cells, rather than the binary system previously used, seems to suggest that audiometric threshold patterns may well be explained by the pattern of hair cell loss and neural loss (P.-Z. Wu et al., 2019; P. Z. Wu et al., 2019). We therefore argue that the classification system, although it has its limitations, remains a good starting point for classification of other forms of hearing impairment, such as genetic forms of hearing loss. It may explain the heterogeneity we see in hearing thresholds based on a more fine-grained picture of the affected structures in the cochlea. The question remains whether such a classification system will ultimately predict the ability to understand speech and speech in noise.

The first study from our group that was published (see Table 1, de Leenheer et al. in 2004) dealt with the effect of mutated *COL11A2* on cochlear function in patients with DFNA13. This deficient gene exhibits a loss of organization of the collagen fibrils in the tectorial membrane, affecting the viscoelastic properties of this membrane. Owing to the near normal CDF and near normal slopes of the loudness growth curves and the tectorial membrane anomaly, it was stated that the hearing impairment acted as a cochlear conductive type of hearing loss. In a second study, similar outcomes (near normal CDF, normal loudness growth) suggestive for a cochlear conductive loss, were reported by Plantinga et al. [2007] in patients with DFNA8/12. These patients also have a disrupted structure of the tectorial membrane matrix due to pathogenic variants in the *TECTA* gene.

Following Schuknecht and Gacek (1993) and Ohlemiller (2004), and based on the outcomes of the psychophysical tests and the present knowledge of the pathology on a cellular level (Nishio et al., 2015), Usher syndrome type 2a, DFNA22, DFNA10 and HDR syndrome have been categorized as ‘sensory’ types of cochlear hearing loss (Leijendeckers et al., 2009; Oonk et al., 2013; van Beelen et al., 2016, 2014) where the hair cells are affected by the specific genetic mutations.

Categorisation of the type of cochlear hearing loss might be complicated by inter-subject variations, as was found in the outcomes of patients with the non-ocular Stickler syndrome (van Beelen et al., 2012). It should be noted that if the deficient gene affects the tectorial membrane, as is the case in non-ocular Stickler syndrome (caused by different pathogenic variants in *COL11A2* than those causing DFNA13), this doesn’t necessarily mean that the hearing loss is uniquely of the cochlear conductive type. Within the group of non-ocular Stickler patients, both sensory and cochlear conductive types of hearing loss seem to be present (van Beelen et al., 2016). The function of the hair cells in these patients might be more negatively influenced by insufficient contact with the impaired structure of the tectorial membrane and by the changes in the elastic properties of the membrane affecting the sensitivity of (otherwise normal) hair cells (Masaki et al., 2009). In contrast to most other groups, this group comprised of patients from different families, what might cause the variation in outcomes.

A limitation of the present retrospective study is the limited number of groups and the limited number of patients per group. Recruiting sufficient numbers of patients with well-established genetic hearing loss is troublesome because most types of genetic hearing loss are rare or very rare. To deal with low numbers, pooling of data from different research centres is important. In addition, it is also recommended to use a universal test battery, like the ‘Auditory profile’ test battery (van Esch et al., 2013; Van Esch and Dreschler, 2015). A second limitation is that although our efforts to limit the range of hearing loss the patients per group were still homogeneous regarding their PTA (i.e. with varying ranges in PTA within the groups, see Table and Figure 2). In contrast to most other studies, the Usher2a and Non–ocular Stickler groups comprised members from different families who had different pathogenic causative variants. The present study suggests that an analysis at family level may provide more information. In addition, previous research from our group has also shown that there is a lot of variation in average sensorineural hearing loss between subjects with specific types of hearing loss (Hartel et al., 2016); the auditory phenotype of patients affected by one single mutation may even vary substantially, and may be caused by modulating variables such as modifying genes, epigenetics and environmental factors (Sadeghi et al., 2013)

Categorization of cochlear hearing loss, e.g. cochlear conductive versus sensory, is important for hearing aid fitting. If the hearing loss is of the cochlear conductive type then linear amplification might be a strategy to evaluate, an approach similar to the ‘classical’ conductive hearing loss. In case of outer hair cell loss, compression amplification might be the better choice to deal with loudness recruitment. Furthermore, in the latter group, noise reduction and speech enhancement might be beneficial to deal with the broader than normal auditory filters (Dillon, 2012). Based on our findings we advise audiologists to fit hearing aids in patients with DFNA8/12 or DFNA13 with a more linear amplification program.

Previously, speech recognition-in-noise scores have often been associated with generic patient data like PTA and age (Pronk et al., 2013; Van Esch and Dreschler, 2015; Vermeire et al., 2016); indeed, present study shows a significant correlation between the speech-in-noise test outcomes and PTA, but not with age. However, regression analysis showed that speech-in-noise and psychophysical data predicted the value of the control subjects within 1 dB, in contrast to an analysis involving CDF and PTA (Figure 2). Although knowing that variables like loudness scaling and DLF are related to hearing loss (Dillon, 2012; Simon and Yund, 1993), the present analysis suggests that psychophysical variables are more sensitive measures of impaired cochlear processing and thus speech recognition than the generic variable hearing loss (PTA), at least at the group-level.

In summary, different types of genetic hearing impairment might uniquely affect cochlear processing, resulting in different auditory profiles, as assessed by psychophysical tests. In the clinic, such knowledge might help to shape the expectations of patients referred for hearing aid fitting. Furthermore, the lack of predictive power at the individual level suggests that there are potentially other variables that could explain more of the variance we observe within the groups. This deep phenotype of hearing loss is needed, if only to have a good tool for selecting the right patients for new and upcoming inner ear therapeutic studies, as an example of precision medicine.

## Data Availability

all data have been obtained from the cited references

